# Application of nasal spray containing dimethyl sulfoxide (DMSO) and ethanol during the COVID-19 pandemic may protect healthcare workers: A randomized controlled trials

**DOI:** 10.1101/2021.07.06.21259749

**Authors:** Ali Hosseinzadeh, Abbas Tavakolian, Vahid Kia, Hossein Ebrahimi, Hossein Sheibani, Ehsan Binesh, Reza Jafari, Seyed Mohammad Mirrezaie, Moslem Jafarisani, Mohammad Hassan Emamian

**Affiliations:** Epidemiology Department, Health School. Shahroud University School of Medical Sciences, Shahroud, Iran; Islamic Azad University, Shahroud Branch, Shahroud, Iran; School of Medicine, Shahroud University School of Medical Sciences, Shahroud, Iran; School of Nursing and Midwifery, Shahroud University School of Medical Sciences, Shahroud, Iran; Clinical Research Development Unit, Imam Hossein Hospital, Shahroud University of Medical Sciences, Shahroud, Iran; Center for Health Related Social and Behavioral Sciences Research, Shahroud University of Medical Sciences, Shahroud, Iran; School of Allied Medicine, Shahroud University School of Medical Sciences, Shahroud, Iran

**Author notes:** Correspond Author: Dr. Moslem Jafarisani, PhD of Clinical Biochemistry, School of Allied Medicine, Shahroud University School of Medical Sciences, Shahroud, Iran. Tell.

**Keywords:** COVID-19, Ethanol, DMSO, healthcare workers, RCT

## Abstract

**Background:** Coronavirus pandemic has affected a large population worldwide. Currently, the standard care for individuals who are exposed is supportive care, symptomatic management, and isolation. The aim of our study was to evaluate effects of combined use of ethanol and DMSO as a nasal spray in preventing COVID-19.

**Methods:** We conducted a randomized controlled trial on volunteer healthcare workers of medical centers that were at the forefront of the fight against COVID-19 in Shahroud, Iran. Two hundred and thirty-two participants were randomly assigned to intervention and control groups to receive DMSO/ethanol or routine care, respectively. The subjects were followed for 4 weeks to determine the incidence of COVID-19 infection in each group based on the RT-qPCR test. Finally, absolute risk difference and relative risk were calculated to evaluate the effect of DMSO in prevent COVID-19.

**Results:** The results showed that the incidence of COVID-19 in the control group and intervention group were 0.07 and 0.008, respectively. The relative risk (RR) was 0.12 (0.9-0.02) according to the incidence rate in the two groups.

**Conclusion:** combined application of DMSO and ethanol in healthcare providers can considerably prevent COVID-19.

## Introduction

SARS-COV-2 is a positive-sense single-stranded RNA virus that causes COVID-19. The coronavirus has affected a large population worldwide since the beginning of the pandemic in December 2019 in Wuhan, China. It has resulted in 155 million infections and 3.24 million death until May 6, 2021(1).

The virus has become a major health challenge to human, health care systems, and global economy (2, 3) for several reasons including (i) the rate, extent, and variety of transmission routes,(ii) the confusing variety of clinical symptoms, and (iii) the unfamiliar and unusual response of the human immune system to COVID-19. Therefore, the current pandemic has become one of the top priorities of all countries (4).

Due to the absence of the virus in bloodstream available antiviral drugs are not effective (5, 6). The production of a potent and stable vaccine against the virus is also questionable because of the instability of the virus genome, which leads to continuous changes in the protein structures of the virus (7-9). In addition, due to technology of production and storage requirements, they are very expensive and their availability is limited especially in developing countries. Therefore, in order to control the disease, preventive methods that are available in all geographical areas are required (10, 11).

SARS-CoV-2 tends to infect the nasal and pharyngeal cavities (upper respiratory tract) for colonization and proliferation for the first few days after entering the body. Therefore, the delivery of effective therapeutics for the prevention and treatment of the disease can be done easily (7-9). The virus only owns the genome and proteins synthesized in the host cell and obtains its phospholipid bilayer from the host cell membrane. Since the virus envelope contains only phospholipids and has no cholesterol, it is very unstable and is a suitable target to destroy the viral particles (12). Thus, hygroscopic and lipolytic agents seem to be able to disrupt the SARS-CoV-2 envelope (13, 14). Such compounds denature proteins structure by absorbing excess water and include (i) Organic solvents such as ethanol, ether, chloroform and DMSO (ii) Ionic and non-ionic detergents such as Tween 80, Triton X100 and SDS (15).

SARS-CoV-2 spike protein plays a crucial role in binding to the angiotensin receptor and entering the lung cell, so unfolding the spike; disrupts this process and the viruses particles cannot continue their life cycle (16, 17). Studies show that ethanol and DMSO have the least toxicity human cells and rats (18, 19). Therefore, using a solution containing 3% dimethyl sulfoxide (DMSO) and 20% ethanol as an intranasal inhalation spray may destroy the viruses’ structure, break the virus transmission chain, and reduce the pathogenicity of the virus.

The present study aims to evaluate effects of a solution of 20% ethanol, 3% DMSO, and 0.1% menthol as a nasal spray in preventing from SARS-CoV-2 infection.

## Methods

A single-center, two-armed 1:1, randomized controlled trial design was conducted to examine effects of DMSO-ethanol nasal spray in the prevention of COVID-19. The study population was healthy personnel of medical centers, who had no history of COVID-19 disease, in Shahroud. Healthy personnel of medical centers in Shahroud (northeast of Iran), who had no history of infection with COVID-19 and volunteered to participate in this trial study, was invited. Two hundred and thirty-two participants were recruited using the convenience sampling method during 2020 from a general referral teaching hospital in Shahroud, Iran (Figure 1). At the beginning of the study, serum IgG and IgM antibodies were tested using a rapid test kit to confirm that volunteers were not infected with COVID-19. The inclusion criteria included all the healthy personnel of the medical centers that were volunteers to participate in the study and those who had a history of seizures or mental illness, allergy to any ingredient included in the spray, acute febrile disease on the day of enrolment, they had received any blood products in the past 4 months; and being unable to comply with the study schedule excluded from the study. The process of conducting the study, the possible benefits and harms of the intervention, the right to withdraw from the study at the desired time, and the confidentiality of the information obtained were explained to eligible individuals and informed consent was obtained from them. The random allocation sequence was created online (www.sealedenvelope.com/simple-randomiser/v1/lists) to randomly determine the composition of blocks (size of each block 4). Then, the participants in the study were randomly allocated to two groups receiving routine care + DMSO combination spray with ethanol and routine care alone (Figure 1). For concealment purposes, the random allocation order was placed in closed opaque envelopes. Due to the nature of the study, it was not possible to blind the participants but the data collector, and the data analyzer were blinded. Enrollment and allocation of participants to study groups was done by a nurse who was trained in this field. Then, a 20 cc spray containing a combination of DMSO and ethanol was given to the individual assigned to the intervention group, and how to use it was taught by the nurse. Individuals assigned to the intervention group were advised to spray a puff of DMSO spray into each nostril every 8 hours for four weeks in addition to routine daily care. It was recommended for the control group to continue daily routine care (Figure 1). The outcome study in the present study was COVID-19 infection. To determine the incidence of COVID-19 in each group, the subjects were followed for 4 weeks. During the follow-up period, RT-qPCR was performed to confirm or rule out the disease in individuals with clinical symptoms of COVID-19. Also, at the end of the fourth week, all participants in the study underwent RT-qPCR to investigate asymptomatic infections. Considering the power of 80%, α=0.05, effect size 0.1, the minimum sample size was estimated to be 93 individuals in each group. The possibility of participant drops out was considered, and therefore the final sample size was determined to be 116 individuals in each group.

**Figure 1.**
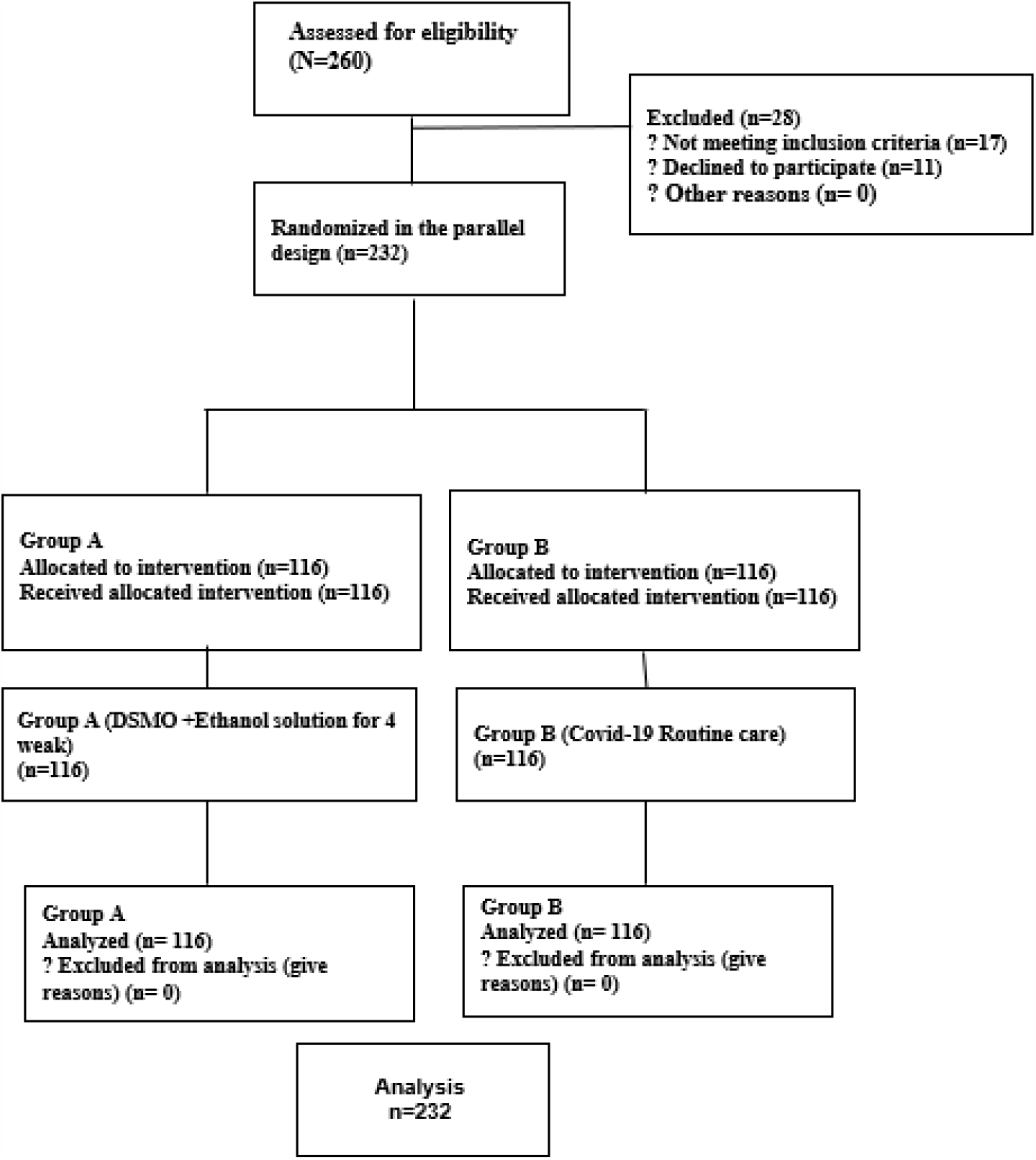
The process of the study according to the CONSORT flow diagram.

The research proposal was approved by the Medical Ethics Committee affiliated with Shahroud University of Medical Sciences (decree code: IR.SHMU.REC.1399.091). The research protocol was registered at the IRCT website under the number of IRCT20200727048217N. We obtained written informed consent from all the participants in 2 copies. In this study, in order to evaluate the effect of using the combination of DMSO with ethanol, absolute risk difference and relative risk were calculated. In calculating the absolute risk difference, the group that received routine care was considered as the exposed group.

STATA version 16 was used for the data analysis. Descriptive statistics including mean and standard deviation for quantitative variables and frequency and percentage for qualitative variables were applied to describe these variables. Statistical tests were two tailed and p-value<0.05 was considered as the level of significance.

## Results

The mean and standard deviation of the participant’s age in this study was 37.18±8.69. Most of the participants 62.3 percent were women and the rest 37.7 percent men. Other characteristics of the participants are listed in Table 1.

**Table 1.** The characteristics of study participants.

| Variable | Intervention group | Control group | P-value |
| --- | --- | --- | --- |
| Age | **$38.28 \pm 9.96$** | **$36.01 \pm 6.96$** | **0.16** |
| Work experience | **$3.82 \pm 1.66$** | **$4.03 \pm 1.67$** | **0.34** |
| sex |  |  |  |
| Men | **48 (40.7)** | **40 (34.5)** | **0.32** |
| women | **68 (59.3)** | **76 (65.5)** |  |
| Residence |  |  | **0.54** |
| Rural | **111 (96.4)** | **5 (3.6)** |  |
| Urban | **113 (98.0)** | **3 (2.0)** |  |

Based on the results of PCR, out of 116 recipients of DMSO-ethanol combination, 1(0.8%) developed COVID-19 during the follow-up period, but out of 116 people assigned to the control group, 8 (6.8%) were diagnosed with COVID-19. The incidence of COVID-19 in the control group and intervention group were 0.07 and 0.008, respectively. According to the incidence rate in the two groups, the relative risk was RR = 0.12 (0.9-0.02). This means that the combination of DMSO with ethanol reduces the risk of COVID-19 by up to 88% (Attributable Fraction Exposer) (Table 2).

**Table 2.** Outcomes of DMSO + Ethanol Solution therapy against COVID-19.

| Intervention | Outcome n (%) |  | Risk | Risk ratio | Absolut risk difference | Attributable fraction exposers |
| --- | --- | --- | --- | --- | --- | --- |
|  | Confirmed COVID-19 | Non confirmed COVID-19 |  |  |  |  |
| **DMSO + Ethanol solution** | 1(0.88) | 115(99.12) | 0.008 | 0.12(0.02-0.97) | 0.06(0.01-0.11) | 0.87 (0.06-0.79) |
| **Routine care** | 8(2.89) | 108(93.1) | 0.07 |  |  |  |

## Discussion

The present study showed that the use of DMSO in combination with ethanol significantly reduced the risk of COVID-19. According to the results, the risk of COVID-19 was about 8-fold higher in those who used routine care than in those who used DMSO spray. Due to the structure of the virus and the mechanism of action of the combination of DMSO with ethanol, the reduction in the risk of COVID-19 in people taking this spray can be justified.

Most viruses including coronavirus species have an envelope, wherein all functionally important virus surface proteins are implanted and secured from dismantling or falling off the virus coat. The virus lipid bilayer is completely derived from host cells, and the virus’s genome has nothing to do with its generation. Therefore, it will not be affected by the extreme mutability of the virus. The physicochemical characteristics of the viral lipid envelope not only help the virus to maintain its spherical structure, but also play a pivotal role as a concrete foundation for the viral surface proteins (20). It is of extreme importance to know that the lipid envelopes of these virus species are highly sensitive to desiccation imposed by hygroscopic chemical agents and lipid-solving substances. Thus, it is utterly simple to conceptualize that even trivial and hardly noticeable changes in the lipid membrane ultra-structural properties could deeply affect the virus infectivity and virulence (21). Considering that most viruses including coronavirus species are wrapped up by an envelope, which is completely derived from the hosting cell, and the virus’s genome has nothing to do with its generation (20).

Therefore, induction of noticeable dryness at the surface of the coronavirus envelope and the nasal cavity epithelial cell membrane as well may cause the process of virus-epithelial cell attachment less.

DMSO is an aprotic polarity solvent that effectively solubilizes a wide variety of organic and inorganic chemicals including lipids with an unquestionable safety profile even at high molar concentrations. It looks as if even low concentrations of DMSO are yet of drastic effect on inducing dehydration and desiccation of lipid membranes. DMSO desiccates and weakens the lipid bio-membrane of cells and microorganisms (22-26). In a comprehensive and professionally designed biochemical study regarding the DMSO inducing dehydration near lipid membrane surfaces C-Y. Cheng concludes that DMSO “sprayed” on lipid surfaces even at low molar concentrations induces profound dehydration of lipid membranes, leading to marked physicochemical changes. He states that the physicochemical effects of DMSO on lipid surfaces are complex and significant at a broad range of DMSO concentrations (27). DMSO is known to directly interact with phospholipid bilayers as well. Molecular dynamics simulations of DMSO-dipalmitoyl phosphatidylcholine systems demonstrate that DMSO modulates the mechanical properties of lipid bilayers, reducing the area compressibility, thickness, solidarity, stability, and bending moduli. These cumulative effects make the lipid layer loose and floppy (28-34). DMSO also causes significant changes on the phospholipid bilayer of cultured skin fibroblast cells and disturbs the quality of the membrane lipid matrix. Numerous biophysical studies clearly demonstrate that the DMSO can induce phospholipid bilayer thinning and create pores through membrane lipid structures. These findings lend support and shed light on the facts behind the antimicrobial and antiviral effects of DMSO. It has been shown that topical DMSO blocks the transcription-replication process of the lipid enveloped, dsDNA containing herpes simplex virus, introducing itself as a direct antiviral agent (35). DMSO has been shown to potentiate the antiviral effects of all disinfectants. DMSO is also among the low toxicity solvents exerting strong free radical scavenger activity. An impressive research work by L. Costa reveals that DMSO decreases cell proliferation and TNF-a, IFNs, and IL-2 production in cultures of peripheral blood lymphocytes. This study signifies the strong anti-inflammatory effects and cytokines storm-preventing capabilities of DMSO (36).

Also, a CDC guideline [2008] regarding chemical disinfectants states that ethanol has generally been underrated as a potent antiviral agent inactivating and disinfecting all “enveloped lipophilic viruses”. The widely accepted explanation for the antiviral action of ethyl alcohol is the denaturation of viral proteins and phospholipid bilayer envelope. In a recent study published by G. Kampf, 80% ethanol was highly effective at killing all 21 tested enveloped viruses within only 30 seconds (37). Another report reveals that ethanol disinfects the lipid-coated viruses instantly in less than 10 seconds. The desiccation and partial denaturation of the virus lipid coat would simply loosen the embedded proteins across the virus envelope. Therefore, it is a reasonable expectation that even low concentrations of a desiccating substance (DMSO) in conjunction with a powerful lipid solvent (ethanol) might execute the desired impact on enveloped viruses. Kampf specifies that the powerful antiviral effects of ethanol were not as remarkable in virus species lacking lipid bilayer coat. It obviously indicates that the primary target for antiviral effects of ethanol is the virus lipid coat (37).

we know that viruses are obligatory and obsessed inhabitant of nasal cavity mucosa, throat, pharynx, and later in its pathogenesis, the lower respiratory tract epithelial cells, alveolar pneumocytes, and capillary endothelial cells (38). We also know that SARS-COV-2 has no noticeable desire to present high viral load viremia (39) and uses angiotensin-converting enzyme-2 (ACE-2) as the major binding site to enter the cells, and we know that the upper and lower respiratory tract harbors the highest concentrations of this protein. Thus, the SARS-CoV-2 must be regarded as a restricted inhabitant of our respiratory tract. Based on these practical reasons, we can constantly intrude and continuously disturb the coronavirus replication milieu in nasal cavities and oropharyngeal area via a nasal spray composed of DMSO and ethanol and causing total turmoil in the breeding and replicating virus herd inside the nasopharynx and throat.

## Strengths and limitations

We acknowledge that this trial has some limitations. First, this phase 3 trial started before the full analysis of the data from the phase 1 and 2 study. Second, it was not possible to blind the study groups. Third, the total dose of iodine absorbed by the suggested regimen is not known exactly. The main strength of this study is, first, DMSO and ethanol have very few contraindications as a mouthwash or nasal spray. second, administration is cheap, simple and rapid. Third, it is readily available in healthcare worldwide. Forth, sensitization is extremely rare.

## Conclusion

The results of this study showed that the use of combination DMSO and ethanol in healthcare providers can considerably prevent COVID-19.

## Recommendation for practice

This study highlighted that the use of DMSO and ethanol is an effective way for the prevent of COVID-19 in medical centers. As a result, the findings of this study if replicated by other studies can be used by healthcare providers, such as physicians, and nurses to reduce risk of COVID-19.

## Data Availability

The Data will available if needed.

## Funding source

This work was supported by Shahroud University of Medical Sciences

## Declaration of competing interests

The authors declare that there is no conflict of interest

## Acknowledgments

This study was supported by grant No 9956 from Shahroud University of Medical Sciences.

## Notes

### Competing Interest Statement

The authors have declared no competing interest.

### Clinical Trial

IRCT20200727048217N

### Clinical Protocols

https://fa.irct.ir/trial/49924

### Author Declarations

We have an ethics committee approval cod from Shahroud University of Medical Sciences with a code of IR.SHMU.REC.1399.091.

### Summary of Updates

Thank you for your contribution, We make a revision and submit it again. Thank you

